# AI-Powered Test Question Generation in Medical Education: The DailyMed Approach

**DOI:** 10.1101/2024.11.11.24317087

**Authors:** J. van Uhm, M.M. van Haelst, P.R. Jansen

## Abstract

**Introduction:** **L**arge language models (LLMs) presents opportunities to improve the efficiency and quality of tools in medical education, such as the generation of multiple-choice questions (MCQs). However, ensuring that these questions are clinically relevant, accurate, and easily accesible and reusable remains challenging. Here, we developed DailyMed, an online automated pipeline using LLMs to generate high-quality medical MCQs.

**Methods:** Our DailyMed pipeline involves several key steps: 1) topic generation, 2) question creation, 3) validation using Semantic Scholar, 4) difficulty grading, 5) iterative improvement of simpler questions, and 6) final human review. The Chain-of-Thought (CoT) prompting technique was applied to enhance LLM reasoning. Three state-of the art LLMs—OpenBioLLM-70B, GPT-4o, and Claude 3.5 Sonnet—were evaluated within the area of clinical genetics, and the generated questions were rated by clinical experts for validity, clarity, originality, relevance, and difficulty.

**Results:** GPT-4o produced the highest-rated questions, excelling in validity, originality, clarity, and relevance. Although OpenBioLLM was more cost-efficient, it consistently scored lower in all categories. GPT-4o also achieved the greatest topic diversity (89.8%), followed by Claude Sonnet (86.9%) and OpenBioLLM (80.0%). In terms of cost and performance, GPT-4o was the most efficient model, with an average cost of $0.51 per quiz and a runtime of 16 seconds per question.

**Conclusions:** Our pipeline provides a scalable, effective and online-accessible solution for generating diverse, clinically relevant MCQs. GPT-4o demonstrated the highest overall performance, making it the preferred model for this task, while OpenBioLLM offers a cost-effective alternative.

## INTRODUCTION

The development of novel educational tools is essential for advancing medical education and finding new ways to challenge the professional growth of clinicians and those in training. Generative artificial intelligence (AI), more recently in the form of large-language models (LLM)^1^, hold promises to positively impact education by its ability to explain complex information, interact with learners to improve learning experiences, and create personalized content that adapts to individual learning levels^1,2^. These models, trained on large input datasets and including over 100 billion parameters^3^, are able to provide trustworthy information and pass professional-level exams in the field of medicine^4^, law^5^ and language^6^. The relative strength of these models can be even further improved by the sequential use of different LLM models, where output from one model can be further improved by a chain of subsequent LLM models^6^. Since LLMs have shown the capability to pass test-questions and provide underlying reasoning for their answers, LLMs may be equally suitable to develop high-quality test-questions themselves and assess the knowledge of others through multiple choice questions (MCQ). Indeed, in recent articles several LLMs models have proven to be useful in MCQ generation in various medical fields^7^, such as pathology^8^ and radiology^9^. The capability of LLMs to generate these questions offers infinite potential for education and training purposes, as the variety of possible questions they can develop is theoretically limitless. Also, question development is flexible and can be responsive to the learner, can be quickly improved, and developed on a very large scale. Currently, many of these AI-driven educational tools lack the ability to generate dynamic, high-quality MCQs that are tailored to specific medical specialties. Also, there is a need for an online and easily accessible platform that leverages LLMs to generate medical test questions, ensuring that these powerful educational tools are easily accessible by a diverse range of users.

In this context, we introduce *DailyMed*, an innovative LLM-based educational tool that can generate test questions such as MCQ across a wide variety of domains within the medical field. We outline the development process of this method, which leverages the capabilities of a number of LLMs. DailyMed combines state-of-the-art artificial intelligence with expert clinical input to create challenging and educational MCQs tailored for medical professionals at various stages of their careers. Furthermore, we evaluate our DailyMed pipeline for questions in the field of clinical genetics, as a benchmark to assess its accuracy and efficacy in producing relevant and reliable questions.

## METHODS

### 1. Quiz generation

DailyMed automatically generates a set of MCQ, covering a wide range of topics within certain medical specialties. The steps in our pipeline are illustrated in **Fig. 1**. The OpenBioLLM model was obtained via the Ollama server (https://ollama.com/taozhiyuai/openbiollm-llama-3/tags), for the other two models we used the API, as these weights have not been publicly released. (**Table 1)**.

**Figure 1.**
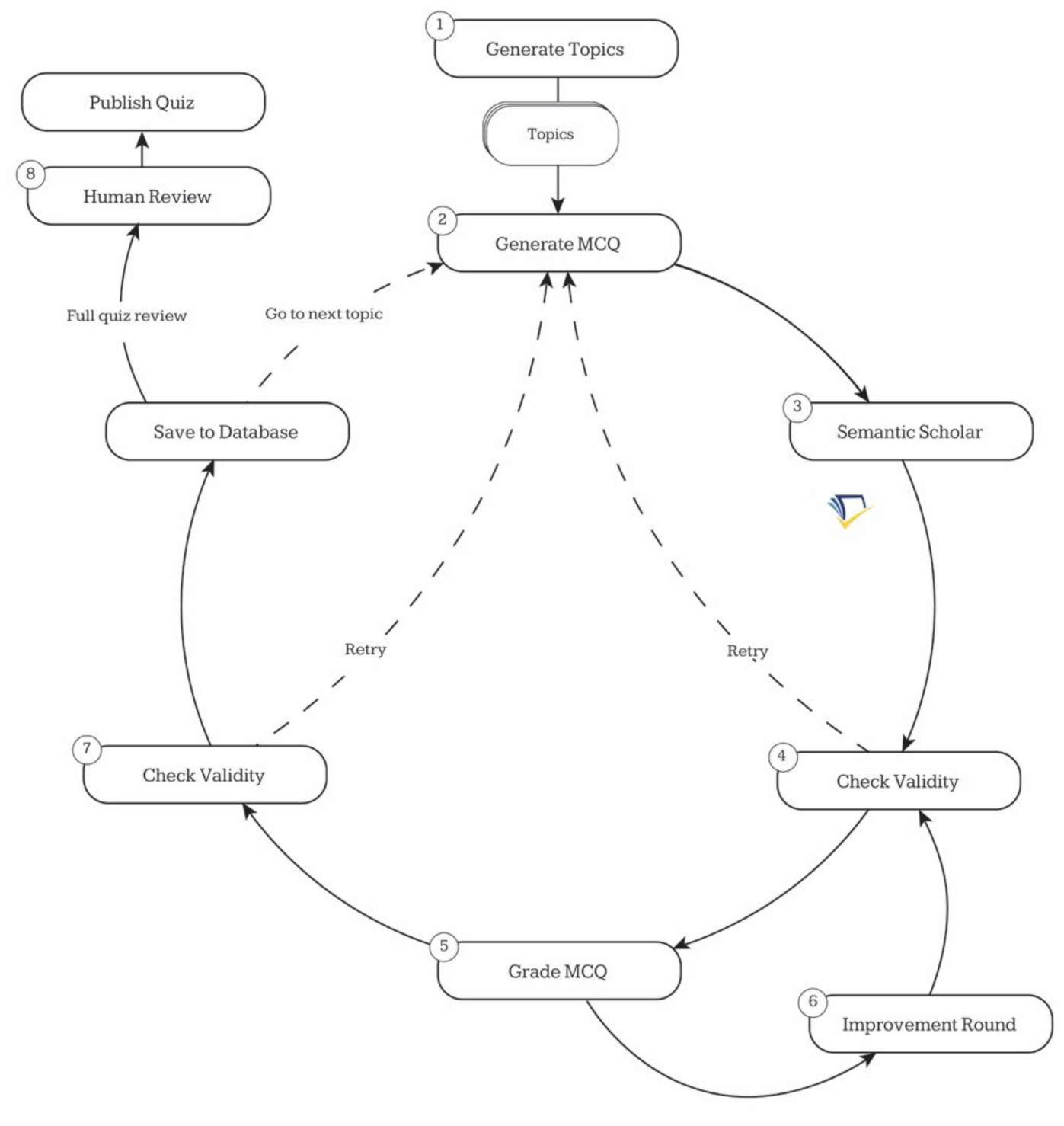
Overview of the Quiz Generation Pipeline from DailyMed. **(1)** Generate a set of 10 topics. **(2)** For each topic generate a question, answer options, a hint, and an explanation. **(3)** Retrieve papers from semantic scholar based on a query generated from the question. **(4)** Use the abstracts retrieved from step 3 to verify the quiz question. If the question could not be verified, the pipeline returns to step 2. **(5)** Grade question based on given criteria. **(6)** Critique the question and apply the critique given on the MCQ. **(7)** Check the validity of the question based on different criteria. If so, the question is saved to the database, and the next question is generated. **(8)** After quiz generation, the quizzes are reviewed before being published. MCQ: Multiple Choice Question.

**Table 1.** Overview of three large-language models that were used in these analyses.

| Model | Source | Inference method | Reference | Number of parameters |
| --- | --- | --- | --- | --- |
| GPT-4o | OpenAI | API | https://platform.openai.com/docs/models | >200 billion |
| OpenBioLLM-70B | Hugging Face (aaditya) | Ollama | https://huggingface.co/aaditya/Llama3-OpenBioLLM-70B | 70 billion |
| Claude 3.5 Sonnet | Anthropic | API | https://www.anthropic.com/news/claude-3-5-sonnet | 175 billion |

The included steps in question generation are as follows:

#### 1.1 Topic generation

The LLM model is prompted to generate a set of 10 topics (**Fig. 1**, Step 1). These topics are based on specific guidelines, such that topics: 1) are specific and focused; 2) cover a wide range of sub-areas within the given subject; 3) test both knowledge and (clinical) application of concepts; 4) are not overly broad or vague; 5) are related to recent developments or current issues in the field; 6) avoid subjective, country-specific or sensitive topics (law, ethics, psychosocial factors).

#### 1.2 Question generation

Results from *query 1.1* are forwarded to the LLM model that iterates over the set of topics (**Fig. 1**, Step 2). For each topic, the model is instructed to provide a question and a set of answering options, a hint which users can help to identify the right answer, and an explanation on why a certain answer is correct, so that a learner can learn from mistakes.

#### 1.3 Semantic Scholar

Questions are checked on validity based on papers retrieved from Semantic Scholar^10^ (**Fig.1**, Step 3). For each question from *1.2 Question Generation* a search query is generated by Claude 3.5 Sonnet. This query is based on the question, correct answer, explanation, and is limited to two concepts of two keywords each (e.g., ‘genetic testing AND Huntington’s disease’). From the top 5 papers (closest vector distance to query), the title, abstract, journal, influential citation count, journal, URL and other identifiers (such as PubMed) are then retrieved and used alongside the question information to let the LLM verify this question based on sources (**Fig. 1**, Step 4). If no supportive literature is found, the pipeline returns to question generation (Step 2).

#### 1.4. Grading

Validated questions are then automatically graded according to difficulty levels: easy, medium, hard, or expert (**Fig. 1**, Step 5). This seeks to evaluate its difficulty level, which are subclassified as ‘easy’ which can be regarded as high-school level knowledge; ‘medium’ which is equivalent to college level; ‘hard’ questions on university level, and ‘expert’ level for questions requiring knowledge typical of an expert or healthcare provider working in the field. If the grade is considered ‘hard’ or ‘expert’, the improvement round (**Fig. 1**, Step 6) is not necessary, and the pipeline will go to *1.5 validation*. We thus chose not to include questions of easy or medium level.

#### 1.5. Improvement round

Questions that are considered easy, or medium are required to go through this improvement process (**Fig.1**, Step 6). Here, the language model evaluates the question and is instructed to provide 3 points for improvement of the question (critique). Improvement ideas include for example:

1. Changing the question format to a case study where a patient presents with symptoms consistent with Duchenne Muscular Dystrophy (DMD), and asking about the likely molecular mechanism underlying their condition.
2. Adding more answer choices that involve other muscular dystrophies or genetic disorders affecting muscle function, requiring the test-taker to differentiate between them based on molecular mechanisms.
3. Asking about the specific location within the gene where pathogenic variants typically occur in DMD, such as ‘What region of the DMD gene is most commonly affected by variants leading to Duchenne Muscular Dystrophy?’

Following this, the critique is piped into the next LLM query, which chooses one of the three ideas given, and incorporates this into the question. The question is then passed back to Step 4. There is a maximum of three attempts to improve the question; if it’s still easy or medium after these, it will be used as is, without further efforts to increase its difficulty.

#### 1.6. Validation

After one or multiple cycles across previous steps, a question is subjected to validation in the last step (**Fig. 1**, Step 7). This validation evaluates whether the question, answer options, hint and explanations are highly likely to be correct. Conflicting and low-confidence questions are being returned to the question generation (step 2). Valid questions are then saved to the DailyMed database (including sources, the un-edited question).

#### 1.7 Human Review and Publishing

Finally, completed quizzes are subjected to human review to ensure quality, accuracy and validity before being published. In this stage, the expert reviewer can change any part of the questions. All these changes are tracked and stored in the database. The end results of these pipeline steps are a high-quality question on a sufficient difficulty level and distinctive answers. The reviewed and published quizzes are available on the DailyMed website and rotated throughout different 10-question quizzes.

### 2. Prompting

In our prompting strategy, we use the Chain-of-Thought (CoT) technique to increase the reasoning capabilities of the LLM during generation pipeline. In Step 3 (Semantic Scholar), the LLM is instructed to evaluate the generated question (1.2) based on the abstracts of the retrieved sources. Here, we let the LLM reason first, before providing an answer (example provided in **Table 2**), allowing us to capture both the reasoning process and the resulting evaluation. This approach enables the LLM to self-assess and refine questions based on its own reasoning, aligning with findings that LLMs can self-improve through self-generated reasoning without extensive external supervision^11^. We use the same CoT technique for 1.4 grading, 1.5 improvement, 1.6 validation, albeit with different reasoning fields and criteria. The applied prompts can be found in the supplementary methods and/or on github (https://github.com/Uhm-J/DailyMed).

**Table 2.** Overview of LLM reasoning output.

| Category | Details |
| --- | --- |
| Verification | The quiz question is verified. DNA methylation is involved in the repression of gene expression. Sources 1 and 4 provide evidence that DNA methylation plays a role in repressing transcription by binding to specific regions of genes, leading to a decrease in their expression. Source 3 also supports this notion by showing that direct inhibition of transcription factor binding is the prevailing mode of gene repression by DNA methylation. Although source 2 does not directly mention DNA methylation's role in gene repression, it discusses MeCP2's function as a transcriptional repressor, which can be linked to DNA methylation. Source 5 further emphasizes the importance of DNA methylation in regulating gene expression and development. |
| Verification Status | PASS |
| Difficulty Reasoning | The question pertains to the role of DNA methylation in gene expression, which is an advanced genetic topic requiring a university level of understanding. |
| Difficulty | Hard |

### 3. Models

OpenBioLLM-70B (8b quantized) (https://huggingface.co/aaditya/Llama3-OpenBioLLM-70B) is a finetuned Llama-3 model on a biomedical dataset. It consists of 70 billion parameters and requires approximately 75 gigabytes of VRAM. To run this, we use RunPod, an external hosting platform. On this platform, it is possible to rent a temporary container (or Pod) with powerful GPUs. Inference is run through Ollama (version 0.3.12). To reduce costs, we do not rent storage containers, but instead download the model upon starting the quiz generation. And generate multiple quizzes sequentially. Additionally, we ran the pipeline with OpenAI’s GPT-4o, and Anthropic’s Claude 3.5 Sonnet, which are two state-of-the-art foundation models (**Table 1**).

### 4. Question evaluation

To assess the validity of our approach to generate valid multiple-choice questions, human ratings by clinical experts are still necessary. We analyzed these questions along several quality criteria:

- *1. Validity*: the question and its proposed correct answer are rated for being factually correct or incorrect as a binary outcome.
- *2. Clarity*: Questions are evaluated whether the question (and answers) is clearly formulated and unambiguous (i.e. not being multi-interpretable) and can be answered from the provided information. *Clarity* is rated on a scale from 1 (=question and or answers are unclear) to 5 (=clearly formulated question and answers).
- *3. Originality*: the question is assessed for originality in its topic, the type of question and the provided set of answers. *Originality* is rated on a scale from 1 (=low creativity) to 5 (=high creativity).
- *4. Relevance*: questions are scored based on their relevance for clinicians and their usefulness in daily practice. Relevance is rated on a scale from 1 (=irrelevant) to 5 (=highly relevant).
- *5. Difficulty*: question difficulty is assessed by the requirement of prior knowledge and experience, understanding of the topic on a deeper level, and distinctiveness between correct and alternative answers (i.e. the presence of ‘distractors’ that are similar to the correct answer). Difficulty is rated on a scale from 1 (=easy) to 5 (=difficult).

## RESULTS

### DailyMed website

The DailyMed website (v1.0) is freely available via https://dailymed.ai/. A user-account is necessary that is able to track progress across different sets of questions and compare results with score results from other users.

### Question analysis

We rated generated questions among five dimensions, including question validity, difficulty, clarity, originality, and clinical relevance, generated by the three different LLM models. Questions were specifically aimed at cases in the field of clinical genetics. Overall, OpenAI’s GPT4-o model showed the highest average scores on all variables **(Fig. 2).**

**Figure 2.**
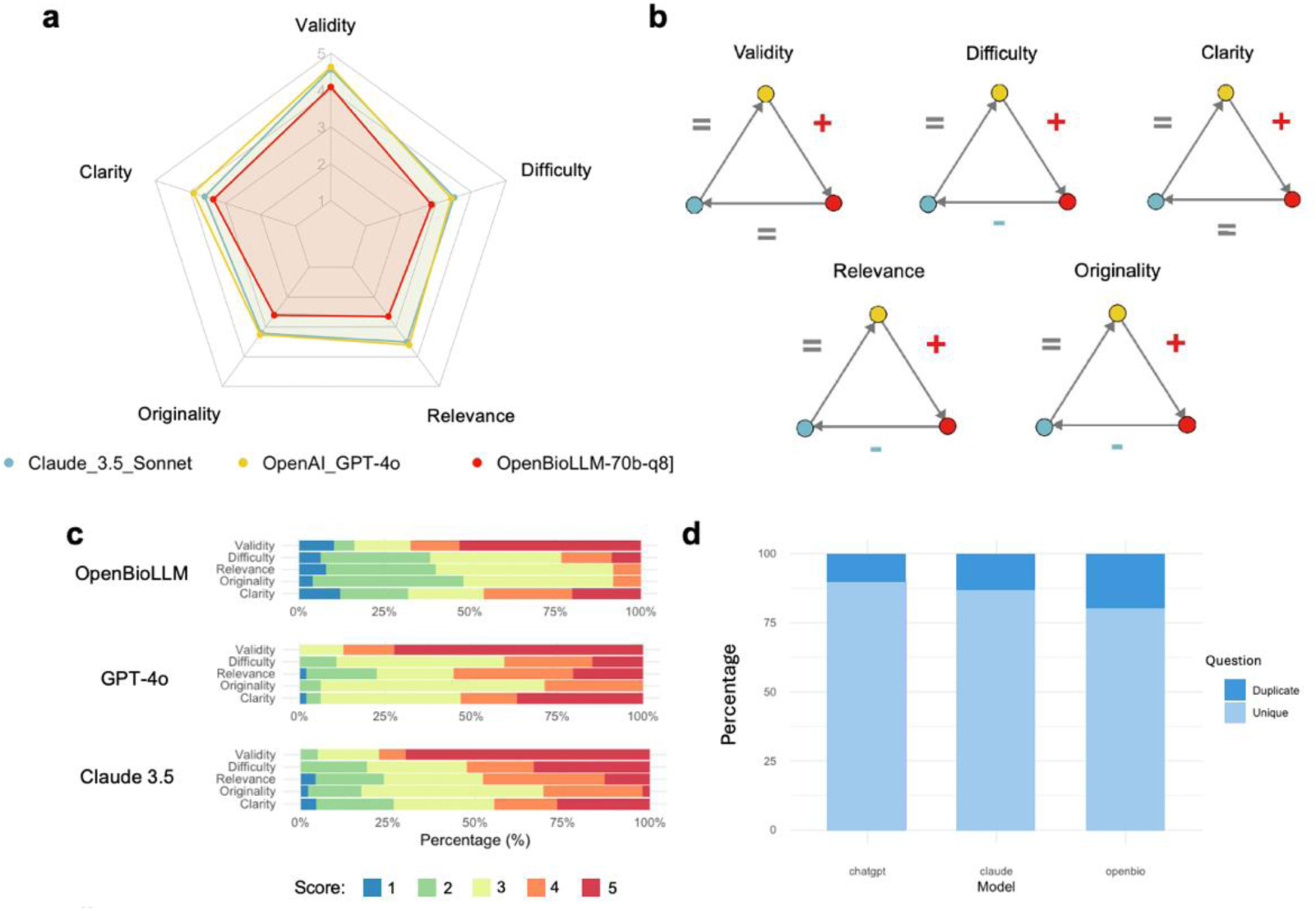
Analyses of multiple choice question generation by the DailyMed pipeline. A) spiderplot showing average scores on each question quality axis for the three tested LLM models; B) plot showing statistical comparisons between these models, where a plus sign represents a higher score, minus sign a lower score, and equal sign an equal score, colored dots represent the different models; C) distribution of quality scores as a percentage of rated questions (n=50 for each model); D) Proportions of unique and duplicate topics covered across quizes

Regarding validity, we observed high validity ratings of questions, answers and rationale generated by the three LLM models, suggesting all three can be used in our pipeline to develop valid questions. The highest validity score was observed by GPT4-o generated quiz questions question (average: 4.6, range: 3-5) compared to Claude (average: 4.4, range: 2-5) and OpenBioLLM (average: 3.9, range: 1-5). Statistical testing showed that the Validity score of OpenBioLLM was significantly lower than that of OpenAI_GPT-4o (mean difference = −0.66, p=0.0082), while no significant differences were found between Claude_3.5 and the other models. On the originality axis, Claude and GPT-4o scored similar (p = 0.78), whereas OpenBioLLM had significantly lower scores compared to both these models (p = 0.0002 and p < 0.001, respectively). OpenBioLLM created significantly easier questions compared to both Claude (p = 0.001) and OpenAI (p = 0.019), with again no differences between these last two models.

Also, for relevance, there was no significant difference scores between Claude_3.5 and GPT-4o (p = 0.64), whereas OpenBioLLM had significantly lower scores compared to the first two models (Claude_3.5, p = 0.0013; OpenAI_GPT-4o p < 0.001). Lastly, questions showed slightly lower clarity in OpenBioLLM-compared to GPT-4o (p = 0.040), but not with the Claude model. As a summary, we found that the GPT-4o model achieves the most valid questions that are also clearly formulated, and rated as more creative, clinically relevant and more difficult than the other models.

### Cross-quiz analysis

In addition to question quality, it is of importance for a multiple-choice quiz to provide sufficient variety in topics. Excessive repetition of the same topics across sessions may limit the learning experience by reducing engagement and hindering exposure to a broader range of concepts. We analysed how often the different models were able to base questions on unique clinical topics (**Fig. 2c**). Here, we found that the GPT-4o model showed the largest diversity (89.8% of questions based on unique topics) compared to Claude (86.9%) and OpenLLM (80.0%).

### Low/high score examples

Examples of a question that was scored relatively high and low respectively are shown in **Fig. 3**.

**Figure 3.**
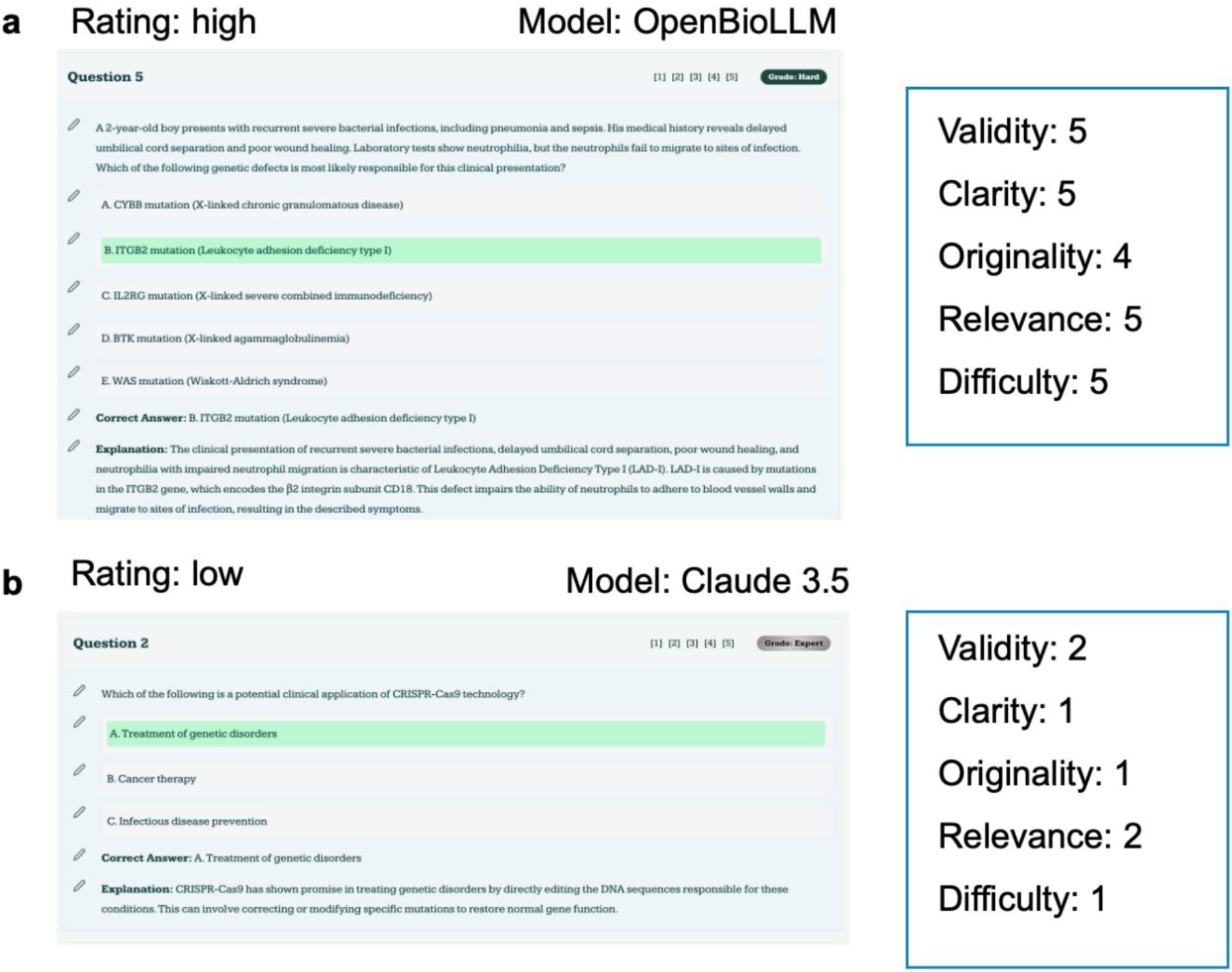
Examples of questions that scored high and low respectively on question ratings. A) example of a highly rated question and rating scores; B) example of a lowly scored question and rating scores.

The first question shows a high score on the five factors (model: GPT-4o). The clinical case description makes the question more engaging and clinically relevant. The text provides clues about the potential differential diagnosis of this clinical presentation. The question formulation is clear and provides several plausible distractors as alternative answers (e.g. the case description concerns a boy and several diseases with an x-linked inheritance are provided). It concerns a relatively rare disease that is not often encountered in the clinic, which contributes to its originality rating. Also, the difficulty of the question is here influenced by the rarity of the disease and the alternative answers. We note that, although the difficulty level is scored higher, this does not mean that it is a better question per se. For a relatively new audience, a somewhat less difficult question may be preferred (e.g. genetic disorders that are more often seen in the clinic, such as Fragile X syndrome, hereditary breast/ovarian cancer, Lynch syndrome).

In contrast, from the lower scored question (model Claude 3.5), it is clear that the topic is (currently) less clinically relevant (CRISPR-CAS gene-editing), as it is not formulated in the context of a clinical problem. There are less provided answers, and we rate these as not being valid. The alternative answer of possibilities for cancer therapy is just as correct as the suggested answer^12,13^. The topic and formulation of the questions are low in their originality, and clarity is low (‘potential clinical application’ is vague and quite broad, anything could have a potential clinical application in some sense).

### Performance and cost-efficiency

The pipeline for generating quizzes using large language models (LLMs) was evaluated in terms of runtime and cost efficiency. **Fig. 4** shows the runtime of the pipeline per question for each model. The cost per quiz varied based on the model used.

**Figure 4.**
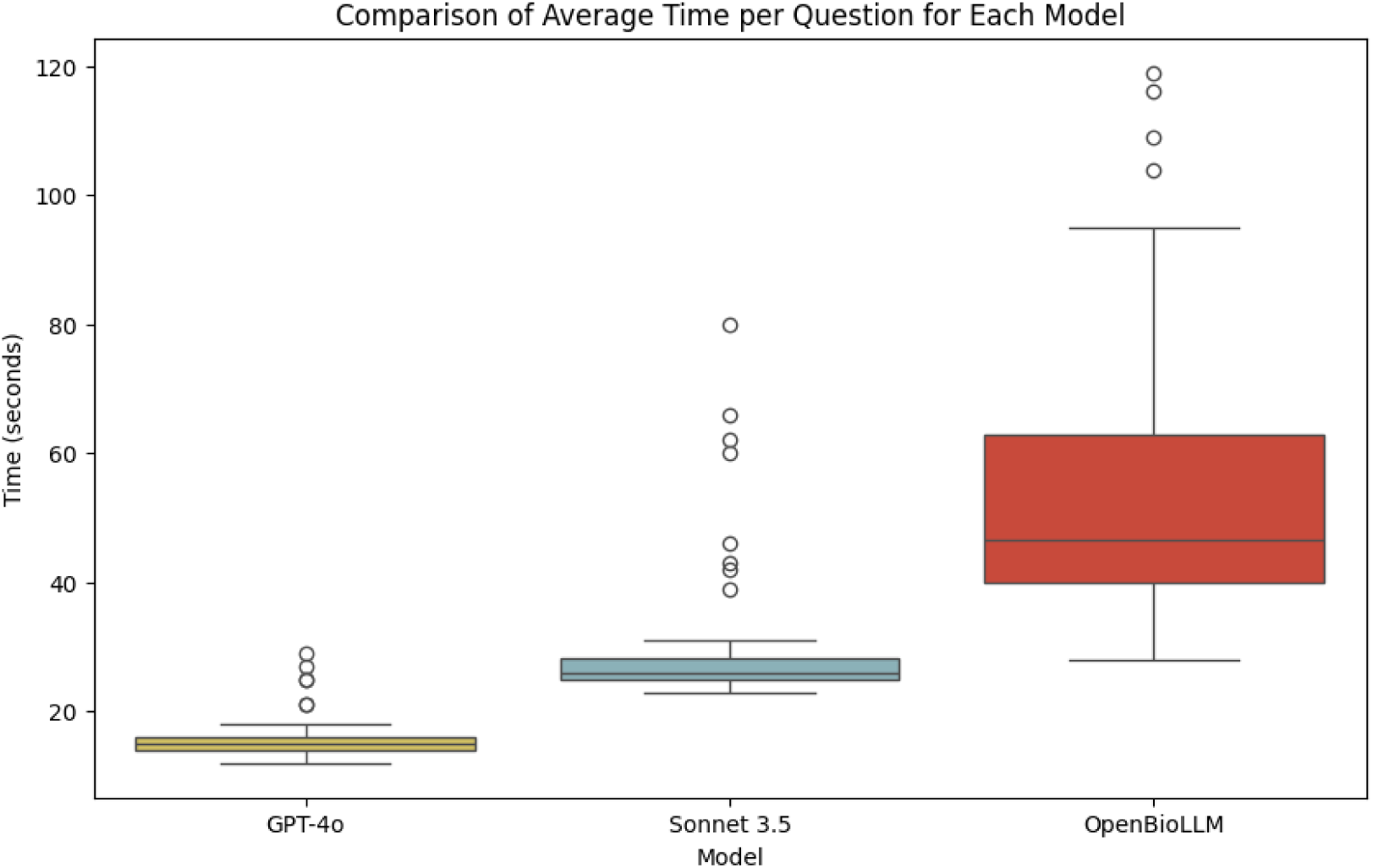
Analyses of runtime per question for the three different models. Boxplot showing the time for question generation, where colors represent three different LLM models.

For OpenAI’s GPT-4o, the average cost per quiz (=10 questions) was $0.51, with an average runtime of 16 seconds per question. For Anthropic’s Claude 3.5 Sonnet, the cost was higher at $0.79 per quiz, with an average runtime of 31 seconds per question. For the OpenBioLLM model, although the average runtime was significantly longer at 53 seconds per question, the overall cost efficiency depended on the number of quizzes generated. The model requires a download time of approximately 25 minutes at the start, translating to a fixed cost of $0.80. After model initialization, the average runtime for a single quiz was 544 seconds, resulting in an average cost of $0.11 per quiz, excluding the model download cost.

## DISCUSSION

In this study, we found that our DailyMed pipeline is able to generate high-quality multiple-choice questions that can be used at scale. Of the state-of-the-art LLM models, we found that for this application in clinical genetics, OpenAI’s GPT-4o model performed best across different quality measures. One intuitive reason for the differences in model performance could be the size of the models. GPT-4o and Claude are significantly larger (with unknown exact parameter counts but speculated to be 200 and 175 billion, respectively). This larger size enables these models to construct more complex sentences with better attention mechanisms, which in turn improves adherence to guidelines and overall quality. Another factor could be better reinforcement learning to improve output preferences. Models like GPT-4o and Claude 3.5 Sonnet likely benefit from more reinforcement learning techniques and data, allowing them to align generated outputs more closely with human preferences, resulting in higher quality multiple-choice questions. At the same time, we observed considerable differences in run time (best model: GPT-4o) and costs (best model: OpenBioLLM) between these models.

As this overview describes the first version of our DailyMed pipeline, we identify several possible improvements. One potential improvement could involve utilizing OpenBioLLM to generate the initial question, followed by GPT-4o mini to refine or rewrite it. This approach leverages the efficiency of OpenBioLLM for question generation while taking advantage of GPT-4o mini’s ability to perform simple rewriting tasks without relying heavily on its extensive knowledge base. This could optimize both performance and resource use.

Furthermore, other subspecialties will be added to the website in addition to clinical genetics, spanning other areas within the medical field (e.g. pediatrics, internal medicine). With the further development of AI and LLM models specifically, regular tuning and updating to the latest LLM models is necessary. The current pipeline will also be explored for additional models that may provide cost-effective or efficient alternatives (such as the Gemini model^14^).

We envision several applications and target audiences for whom our online tool can provide a useful learning experience. First, medical student with an interest in certain medical specialties, such as clinical genetics, can get more familiar with different case presentations in the clinic and the decision-making process in these cases (e.g. before starting a residency in this subspecialty). With an estimated 45 unique topics (89% for GPT-4o) that are found in 5 quizzes (50 questions), a student that is not familiar with genetics may thus encounter 180 topics in a month’ time with only a couple of minutes of daily practice and reflection. Secondly, coordinators of residency programs for clinical genetics residencies may incorporate quiz questions in their curriculum, either as a formal test, or as a way of self-reflection on possible knowledge gaps. Given limited time that residents may have for self-study in between clinical duties, an automated approach of using LLM for topic and question generation may be a valuable solution that allows residents to focus on high-yield content while minimizing the time spent on seeking new information. Finally, experienced clinicians can use DailyMed to stay updated on evolving knowledge, particularly in rapidly advancing fields like clinical genetics. Additionally, this tool can support continuing medical education (CME) initiatives, enabling clinicians to efficiently assess their knowledge in areas where they may need review.

LLM models continue to develop rapidly, as illustrated by the number of parameters included in the latest models (>200 billion in GPT-4o). We envision that such models may become more responsive to the learner’s progress and can be continuously updated to provide new material and/or new areas in which the learner can develop. Also, as generative AI and advanced LLMs allow the generation of images from text^15^, or even video^16^, multimedia material may be integrated in multiple choice questions, providing a more dynamic and interactive learning experience. Examples could be the presentation of certain clinical symptoms, dysmorphological features or congenital anomalies (which would not influence a patient’s privacy) and visualization of imaging results, anatomical diagrams, or 3D reconstructions.

To conclude, our pipeline demonstrates the potential of large language models, particularly GPT-4o, to efficiently generate high-quality multiple-choice questions for medical education. Future versions of this system could integrate more specialized clinical input and explore additional models to further optimize question diversity, accuracy, and accessibility for a broader range of medical professionals.

## Data Availability

All data produced are available online at dailymed.ai/paper & github.com/Uhm-J/dailymed

https://dailymed.ai/paper

